# Segpy: a streamlined, user-friendly pipeline for variant segregation analysis

**DOI:** 10.1101/2024.12.26.24319616

**Authors:** Michael R. Fiorini, Saeid Amiri, Allison A. Dilliott, Dan Spiegelman, Guy Rouleau, Sali M.K. Farhan

**Author notes:** Corresponding author: Sali M.K. Farhan.

## Abstract

Understanding the role of genetic variants in disease is essential for diagnostics and the advancement of genomic medicine. While the advent of high-throughput sequencing has been matched by the development of sophisticated genomic analysis tools, these packages often involve complex analytical procedures that can be challenging for researchers with limited computational experience. Additionally, modern genomic datasets require high-performance computing (HPC) systems, which may be difficult to implement for unfamiliar users. To address these challenges, we introduce Segpy, a streamlined, user-friendly pipeline for variant segregation analysis that integrates seamlessly with HPC environments. Segpy supports single-family, multi-family, and population-based datasets, allowing researchers to evaluate how genetic variants co-segregate with disease in pedigree-based analyses and compare allele frequencies between affected and unaffected individuals in case-control analyses. To date, the application of Segpy has facilitated the identification of genetic variants contributing to many human diseases and is now available as a publicly available framework.

## Introduction

Traditional pedigree-based analyses, including family trios, quartets, and multi-generational families, have been key to characterizing Mendelian diseases (1). Specifically, segregation analyses that examine how genetic variants co-segregate with disease can assess inheritance patterns, zygosity, potential risks for future generations, and diagnostics, while also nominating possible genetic underpinnings of disease. However, pedigree-based methods are less effective for studying complex diseases due to the potential involvement of multiple genetic factors with small effect sizes (2). In contrast, population-based case-control cohorts, which compare allele frequencies between large groups of affected and unaffected individuals, are sufficiently powered to detect common alleles of small effect through genome-wide association studies, while also identifying rare variants through gene burden analyses (3). Both pedigree-based and population-based studies rely on accurate imputation of allelic counts at millions of variant sites in affected and unaffected individuals, which can present significant challenges for investigators with limited computational expertise.

Advancements in understanding the genetic determinants of disease have been driven largely by improvements in high-throughput DNA sequencing technologies, enabling whole-genome and whole-exome sequencing (WGS/WES). Naturally, the rise of high-throughput sequencing technologies has corresponded with the development of sophisticated genomic analysis tools (4-6). However, these comprehensive packages often constitute complex installation, configuration, and execution requirements, posing a steep learning curve for researchers with limited computational experience. Additionally, the size of modern genomic datasets requires high-performance computing (HPC) systems to meet the substantial data processing and storage demands, which can be challenging to implement for unfamiliar investigators. To address these issues, we introduce Segpy—a streamlined, user-friendly pipeline designed for variant segregation analysis, which integrates directly with HPC environments and facilitates the analysis of single-family, multi-family, and population-based datasets.

### Segpy overview

The Segpy pipeline is designed for variant segregation analysis applied to both pedigree-based family cohorts — those involving single or multi-family trios, quartets, or extended families — and population-based case-control cohorts to compute allelic carrier counts at variant sites across study subjects. As input, users must provide a single Variant Call Format (VCF) file describing the genetic variants of all study subjects, which can be optionally annotated with Ensembl Variant Effect Predictor (VEP) (7) to prioritize genetic determinants, as well as a pedigree file describing the familial relationships among those individuals (if applicable) and their disease status. As output, Segpy computes variant counts for affected and unaffected individuals, both within and outside of families, by categorizing wild-type individuals, heterozygous carriers, and homozygous carriers at specific loci. These counts are organized into a comprehensive data frame, with each row representing a single variant and labeled with the Sample IDs of the corresponding carriers. The data frame serves as the foundation for downstream statistical analyses tailored to the user’s specific research question, including the investigation of variant inheritance patterns, variant co-segregation with disease, and the identification of *de novo* pathogenic variants for pedigree-based analyses, as well as the assessment of allelic frequencies in both affected and unaffected individuals for population-based case-control analyses.

Packaged as a containerized pipeline, Segpy includes all necessary code and software dependencies, simplifying the installation process and standardizing analyses across independent research groups. While implemented as a command-line tool, Segpy was developed with varying levels of computational expertise in mind, prioritizing a streamlined process for investigators. Considering the scale of modern WES and WGS datasets and the computational power required for their analyses, the Segpy pipeline was designed for integration with the users’ HPC clusters, effectively eliminating size limitations on the datasets that can be analyzed.

### Underlying framework

The Segpy pipeline is packaged as an *Apptainer* (v1.2.4) container, offering a standardized environment that encapsulates all required software and dependencies, ensuring compatibility with all Linux systems (8). The containerized framework consists of three main components: (i) analytical scripts written in Python (v3.10.2), (ii) customizable parameter and configuration files, and (iii) job submission scripts written in Bash (**Figure 1**). Users can customize their analysis parameters using the provided configuration file, where they must specify their reference genome build, indicate whether to retain VEP annotations, and choose to utilize a HPC cluster or a local workstation. In the case where users opt to use a HPC cluster, Bash scripts are submitted to create ‘Jobs’—resource requests that depend on the parameters specified in the configuration file, such as CPUs, memory, and time. These jobs are submitted to the HPC system through the *SLURM* scheduler to run the analytical Python scripts (9). To meet the requirements of various study designs, the pipeline integrates the ability to analyze pedigree-based and population-based datasets by providing three distinct, yet highly comparable analysis tracks: 1) single-family, 2) multi-family, and 3) case-control.

**Figure 1.**
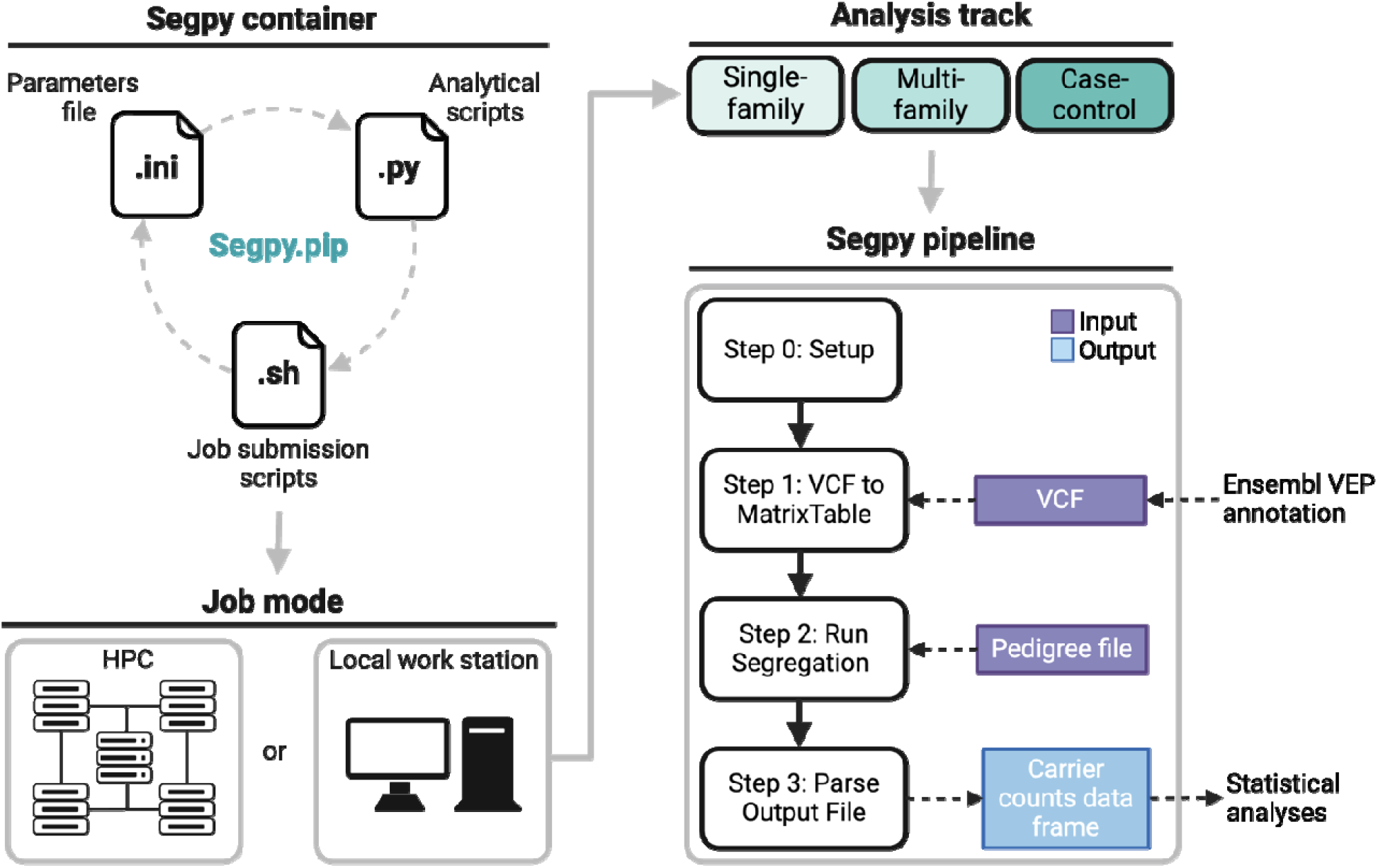
Segpy framework overview. The Segpy container encapsulates all necessary code and software dependencies and comprises three major components: 1) adjustable parameter and configuration files, 2) job submission scripts for high performance (HPC) environments, and analytical Python scripts. Upon installing the Segpy container, users must define their job mode, electing to use either their HPC system or local workstation. Next, users select their analysi track. To accommodate the diverse requirements of various study designs, Segpy offers three distinct, yet highly comparable analysis tracks: 1) single-family, 2) multi-family, or 3) case-control. All three analysis tracks follow a four-step pipeline. In Step 0, a working directory is created to store configuration files and the outputs of subsequent steps. In Step 1, the user-provided Variant Call Format (VCF) file is converted to the *Hail* MatrixTable format to efficiently manipulate large scale datasets. In Step 2, variant segregation analysis is performed based on the user-provided pedigree file to generate a comprehensive data frame that details the number of variant carriers according to disease status and familial status, when applicable. In Step 3, the carrier counts data frame is parsed based on user specifications to reduce the computational burden of analyzing the final dataset. Abbreviations: VEP, Variant Effect Predictor.

Each of Segpy’s three analytic tracks follow a four-step pipeline: Step 0: Pipeline Setup, Step 1: VCF to MatrixTable, Step 2: Segregation Analysis, and Step 3: Parse Output File. In Step 0, a working directory is created for the analysis, where a modifiable configuration file and a text file documenting the job settings and user-defined analytical parameters are stored to ensure replicability. The outputs from each subsequent analytical step are saved in this working directory. In Step 1, the user-provided VCF file is converted to the *Hail* MatrixTable format, which is optimized for efficiently storing and manipulating large-scale genomic datasets (https://github.com/hail-is/hail). All intermediate MatrixTable files are saved in the working directory for easy access and further analysis. In Step 2, variant segregation analysis is conducted based on the user-provided pedigree file, which defines the Sample IDs, disease status, and familial relationships (where applicable) of the study subjects, alongside the outputs from Step 1 to generate a comprehensive data frame that details the number of variant carriers stratified by disease and familial status, when applicable. For pedigree-based analyses, Segpy iterates through each user-defined family to compute the number of wild-type individuals, heterozygous carriers, and homozygous carriers for a given allele at a specific locus, both within the corresponding family and in the broader cohort. For the case-control analysis, the same process is applied, but without the family-wise iteration, to calculate the number of affected and unaffected allele carriers at the given locus. Importantly, the carrier counts are accompanied by the Sample IDs of heterozygous and homozygous carriers to facilitate downstream analyses. In Step 3, the carrier counts data frame is parsed based on user specifications to reduce the computational burden of analyzing the final dataset. Namely, users can eliminate duplicated variant entries from divergent VEP annotations or remove uninformative characters (e.g., quotes and brackets). The underlying code for Segpy is available on GitHub (https://github.com/neurobioinfo/segpy).

### Running Segpy

A comprehensive user guide for running Segpy is available on GitHub (<u>neurobioinfo.github.io/segpy/</u>). The containerized pipeline can be freely downloaded from Zenodo (https://zenodo.org/records/14503733). After installation, users can initiate the pipeline by running Step 0, specifying their desired job mode — “slurm” for HPC clusters or “local” for local workstations — and analytical track — “single_family”, “multi_family”, or “case_control”. The pipeline can be initiated using the following command in the terminal:

~~~
bash segpy.pip/launch_segpy.sh \
~~~

~~~
-d /path/to/working/directory \
~~~

~~~
--steps 0 \
~~~

~~~
--jobmode [slurm | local]
~~~

~~~
--analysis_mode [single_family | multiple_family | case-control]
~~~

Users can then execute the full segregation analysis with the following command:

~~~
bash segpy.pip/launch_segpy.sh \
~~~

~~~
-d $PWD \
~~~

~~~
--steps 1-3 \
~~~

~~~
--vcf $VCF \
~~~

~~~
--ped $PED \
~~~

~~~
--parser [general | unique]
~~~

### Application cases

Despite the vast diversity of cohort configurations and research questions that accompany genetic analyses, the generalizable framework of the Segpy pipeline can be applied to an array of study designs to enable meaningful insights into the genetic underpinnings of disease. This adaptability and capacity to streamline complex analyses has made the Segpy pipeline a cornerstone for genetic analysis among our group and others at the Montreal Neurological Institute-Hospital (The Neuro) over the past decade. Specifically, we have applied this tool in our work, which resulted in >60 manuscripts over the past decade.

Within the pedigree-based analysis track, the Segpy pipeline can accommodate a wide range of inputs, including single-family trios or quartets, multigenerational families, and multi-family pedigrees. Single-family, trio-based segregation analyses have been instrumental in identifying genetic drivers of diseases (1), leading to a tenfold reduction in candidate variants and a 50% increase in diagnostic yield compared to singleton sequencing (10). Moreover, multigenerational and extended families, which include a larger number of affected and unaffected individuals, provide valuable genetic context that enhances the detection of rare causal variants and supports more robust co-segregation analyses (11). Indeed, single-family analyses have proven opportunistic at The Neuro as Segpy has been effectively applied to uncover genetic contributors to a range of diseases, including Pelizaeus-Merzbacher disease (12), hereditary spastic paraplegia (13), and essential tremor (14).

Incorporating multiple families with probands exhibiting similar clinical presentations — whether for an unrecognized rare disease or cases of the same condition — further strengthens evidence for a variant’s involvement in disease causation through consistent co-segregation. Segpy has also been extensively applied in multi-family studies investigating a range of neurological phenotypes, such as autism spectrum disorder (15), bipolar disorder (16, 17), schizophrenia (18), spastic paraplegia (19, 20), and restless leg syndrome (21, 22), among others.

Beyond identifying causal variants and enhancing diagnostic yield, pedigree-based designs are vital for establishing inheritance patterns and assessing risks for current and future family members. They allow for the mapping of Mendelian inheritance patterns, including autosomal dominant, autosomal recessive, and X-linked patterns, while also enabling the filtering of variants with Mendelian inconsistencies. For example, Segpy has been applied to unravel genetic drivers of autosomal recessive hereditary spastic paraplegia (23, 24) and has identified recessive *CDK5RAP2* variants in patients with isolated agenesis of the corpus callosum (25). Additionally, pedigree-based analyses facilitated by Segpy have detected *de novo* variants contributing to disorders such as obsessive-compulsive disorder (26), schizophrenia (27, 28), and developmental and epileptic encephalopathies (29).

In turn, Segpy’s case-control analysis track is specifically designed for population-based cohorts, providing an efficient approach for studying well-characterized phenotypes without the need to enroll family members of affected individuals. While case-control studies are not intended to assess inheritance patterns, they offer a powerful framework for identifying novel disease-associated loci through a variety of downstream statistical analyses. The output from the Segpy pipeline supports multiple analytical approaches, including genome-wide and exome-wide association studies to identify common genetic variants linked to disease, rare variant burden analysis to assess the cumulative impact of rare variants across the cohort, and polygenic risk scoring to evaluate the combined effects of multiple genetic variants on disease susceptibility. Previous case-control analyses using Segpy have implicated rare variants in several diseases, including essential tremor (30), Parkinson’s disease (31-34), rapid eye movement (REM) sleep behavior disorder (35, 36), and congenital hypothyroidism (37). Additionally, the pipeline has been used to evaluate polygenic risk in amyotrophic lateral sclerosis (ALS) (38).

## Conclusion

The Segpy pipeline offers a versatile and accessible solution for variant segregation analysis across diverse genomic study designs, including both pedigree-based and population-based cohorts. Notably, its direct integration with HPC environments enhances analytical efficiency by enabling rapid data processing, effectively eliminating size limitations associated with large-scale datasets. While multiple genomic analysis packages catering primarily to expert geneticists have been developed, their complex frameworks can be challenging for labs without specialized genetic expertise looking to supplement their broad research program (4-6). In contrast, Segpy’s targeted focus on segregation analysis, user-friendly design, and containerized framework empowers researchers to conduct sophisticated analyses without demanding intensive learning efforts to familiarize with the framework. To date, Segpy has proven to be exceptionally valuable at The Neuro in identifying genetic contributors to wide range of neurologic disease from both pedigree-based and population-based study designs. We anticipate that our publicly available framework will expand our in-house success across multiple research groups and institutions, ultimately fostering contributions from investigators with diverse research backgrounds to the growing wealth of genetic discovery to advance the promise of genomic medicine.

## Data availability

Data sharing is not applicable to this article as no datasets were generated or analysed during the current study. The code underlying the Segpy pipeline is publicly available on GitHub (https://github.com/neurobioinfo/segpy).

## Conflicts of interest

The authors declare no conflicts of interest.

## Funding statement

This work was supported in part by grants from CIHR, Brain Canada, and ALS Canada.

## Acknowledgments

The authors have no acknowledgements to declare.

## Notes

### Competing Interest Statement

The authors have declared no competing interest.

